# ‘*Involve those, who are managing these outbreaks*’– Identifying barriers and facilitators to the implementation of clinical management guidelines for High-Consequence Infectious Diseases in Uganda

**DOI:** 10.1101/2024.01.20.24301549

**Authors:** Olive Kabajaasi, Stefan Schilling, Mathias Akugizibwe, Peter Horby, Peter Hart, Louise Sigfrid, Shevin T. Jacob

**Affiliations:** Walimu, Kampala, Uganda; Psychology Department, University of Exeter, Exeter, UK; Pandemic Sciences Institute, University of Oxford, Oxford, UK; Social Sciences Department, Medical Research Council, Entebbe, Uganda; Wellcome Trust, London, UK; Department of Clinical Sciences, Liverpool School of Tropical Medicine, Liverpool, UK

**Keywords:** epidemics, pandemics, High Consequence Infectious Diseases, clinical management guidelines, Filovirus disease, COVID-19

## Abstract

Prior research highlighting the complexity of clinical management guidelines (CMG) implementation, has suggested that limited access to treatments and equipment [1] and substantial issues regarding availability, inclusivity, quality, and applicability [2–6] hinder the implementation of CMGs in Low- and Middle-Income Countries (LMICs). This in-depth case study of Uganda – coincidentally occurring during the 2022 Sudan Virus Disease outbreak – aimed to explore contextual and supplementary factors which hinder or facilitate CMG development and implementation. Using thematic network analysis [7–9] the research describes five thematic topics, that emerged from interviews with 43 healthcare personnel, as barriers to the implementation of CMGs in Uganda, namely: (1) deficient content and slow updates of CMGs; (2) limited pandemic preparedness and response infrastructure; (3) slow dissemination and lack of training; (4) scarce resources and healthcare disparities and (5) patient outcomes. The study displays how insufficient training, patchy dissemination and slow updating exacerbate many of the underlying difficulties in LMIC contexts, by illustrating how these issues are related to resource constraints, healthcare disparities, and limited surveillance and referral infrastructure. Key recommendations to enhance CMG implementation are provided, underscoring the necessity of integrating local stakeholders to ensure guidelines are reflective of the reality of the local health system, applicable and inclusive of resource-constrained settings, available as “living guidance” that is disseminated widely and supported by cascading hands-on training. Findings offer valuable insights for LMICs to improve high consequence infectious disease outbreak responses and for organizations involved in guideline development and funding.

## INTRODUCTION

The COVID-19 pandemic, which resulted in over 700 million people infected worldwide and over six million deaths, highlights our vulnerability to emerging pathogens and the necessity for harmonized guidance for supportive care of critically ill patients, particularly in the early stages of an outbreak of High Consequence Infectious Diseases (HCID) [10]. In the absence of vaccines or directed treatment for most priority pathogens on the World Health Organization (WHO) Research and Development (R&D) Blueprint list [11,12], early and well monitored supportive care is often the only available treatment. In such situations, clinical management guidelines (CMGs) are key tools to maximise patient outcomes by providing clinicians with a standardised approach to administering supportive care and therapeutics for HCIDs [13]. High quality CMGs – accessible by front-line clinicians and inclusive of vulnerable population groups – are crucial for the standardisation of supportive care, in a way that optimises outcomes for patients and safety for healthcare personnel (HCP) [14,15]. Considering the limited empirical knowledge on emerging pathogens, CMGs for HCIDs need to be responsive to incorporating new evidence and disseminating evidence to frontline HCP. Accordingly, during the COVID-19 pandemic, the WHO produced “living” guidance for use by frontline clinicians [16].

However, CMGs produced during previous public health emergencies often did not meet the agreed upon gold standards for CMG development [17,18]. Furthermore, despite the availability of frameworks and guidance for the development of quality CMGs, their development is often impeded by a lack of collaboration and consensus, with checklists for reporting quality often lacking comprehensiveness and contextual relevance [19–21]. For example, during the 2013–2016 West African Ebola virus disease outbreak, it was found that lack of CMGs hindered the standardization of best practices [22] due to limited consensus by response stakeholders for several high-priority supportive care interventions [23]. Recent studies suggest that policy, organisational and inter-personal factors, alongside time and resource constraints (e.g., financial, human and expertise) hinder CMG implementation [24,25]. Similarly, issues appear to exist around HCP awareness of existing guidelines and recommendations [26]. For example, a recent survey of 76 clinicians from 27 countries conducted during the COVID-19 pandemic identified issues in the implementation of COVID-19 CMGs, including limited access to treatments and equipment and lack of guidance for at-risk populations and resource-constrained settings [1]. These findings are supported by a series of systematic evidence reviews of existing CMGs, for viral haemorrhagic fevers (VHFs), chikungunya, mpox and severe acute respiratory syndrome (SARS), which found similar issues regarding availability, inclusivity, quality, and applicability of CMGs [2–6], particularly in Low- and Middle-Income Countries (LMICs).

Considering the difficulties for CMG development and implementation in LMIC settings, this study aimed to assess barriers and facilitators to the adaptation, adoption, and implementation of CMGs for the treatment of Filovirus disease (FVD) and COVID-19, using Uganda as a case study. As a result of multiple FVD outbreaks since 2000 (see Table 1 compiled from [27–30]), the Ugandan Ministry of Health (MoH) developed a pocket manual for frontline HCP comprising guidance to provide optimal and safe management of patients with FVDs, which was subsequently adopted as the basis for WHO guidance for frontline HCP during the 2013-2016 EVD outbreak in West Africa. This history of disease outbreaks and Uganda’s role in the development of supportive care guidance over the last two decades contribute to a rich case study, allowing insight into the process and uptake of CMGs.

**Table 1:** Overview of HCID outbreaks in Uganda with data from [27–30].

| Year | Cases | Deaths | Case fatality rates | High Consequence Infectious Disease (HCID) |
| --- | --- | --- | --- | --- |
| 2000 | 425 | 224 | 53% | Sudan virus disease (SVD) |
| 2007 | 149 | 37 | 25% | Bundibugyo virus disease (BVD) |
| 2012 | 06 | 03 | 50% | Sudan virus disease (SVD) |
| 2012 | 07 | 004 | 36% | Sudan virus disease (SVD) |
| 2007-2018 | 23 | 09 | 39% | Marburg virus disease (MVD) |
| 2020-2023 | 171 871 | 3,632 | 2% | COVID-19 |
| 2022-2023 | 142 | 55 | 39% | Sudan virus disease (SVD) |

The present study aimed to explore 1) contextual factors and processes that shape CMG development and implementation in Uganda and 2) supplementary factors influencing use of guidelines in practice. As such, the study assessed when and why guidelines were mobilised or foregone; how CMGs may be communicated to patients and the public to bolster confidence and trust and how far guidelines accommodate different aspects and priorities of care. This case study was conducted as part of the ‘Evaluation of Clinical Management Guidelines for High Consequence Infectious Diseases’ (ESHCID), which comprised two other case studies and a series of reviews exploring availability, inclusivity, scope and quality of CMGs for HCIDs [2–6].

## METHODOLOGY

This qualitative in-depth case study examined the barriers and facilitators of CMG development and implementation within the Ugandan context. The study results are presented using the Consolidated Guidelines for Reporting Qualitative Research (COREQ) [31].

### Sampling and recruitment

Prior to enrolment, stakeholder mapping was conducted to categorize participants at different levels (i.e., international, national, and local), taking into consideration their previous or current role in the development or adaptation of CMGs. Participants were eligible to participate if they were previously or currently involved in outbreak response or CMG development for FVDs and/or COVID-19. A total of 62 potential study participants from five general and nine regional referral hospitals, MoH headquarters and WHO country office in Uganda were contacted using purposive and snowball sampling techniques (see supplementary information). Of these, 19 declined participation citing urgent involvement in the 2022-23 SVD outbreak response at the time of data collection as reasons.

### Data Collection

Forty-three interviews were conducted between August and December 2022 in English by a female (OK) and a male (MA) social scientist (M.A, Sociology), with ample experience in conducting qualitative research in healthcare settings in Uganda and with no prior relationship with participants. Participants were interviewed online or in private rooms within their respective workplaces, with interviews lasting between 25 to 120 minutes. No other person was present. Unpiloted semi-structured interview topic guide developed collaboratively by the research teams based in Uganda and the United Kingdom was used, with topics focusing on knowledge, development, availability and access, adaptability, inclusivity, and supplementary factors affecting implementation of CMGs (see supplementary information). With the onset of the SVD outbreak in September 2022, the topic guide was revised to include specific questions that captured information on how CMGs were applied in real-time until data saturation was reached, but no follow up interviews were conducted. With permission, interviews were digitally recorded and transcribed verbatim and no participant feedback was collected.

### Analysis

Transcripts were evaluated through thematic network analysis which involved: 1) data familiarization from field notes and raw data; 2) theme identification; 3) data coding and 4) organization of codes and themes and 5) exploration via social network modelling. Two researchers (OK and SS) coded the data using NVivo 12 plus and NVivo 20, utilising a multi-theme coding method developed by the author [8,9] with discrepancies resolved through consensus. Codes were extracted from NVivo, transformed, and imported into a social network programme (Gephi 0.9.5) to develop a visual representation of both the relationships between thematic codes and their importance in the overall network in line with prior network representation of qualitative data [7–9,32,33]. Edge weights and the association measure ‘lift’ were utilised using the elbow method to minimize random links between codes [34–36].

Makerere School of Public Health Research and Ethics Committee (SPH-2022-265), Uganda National Council for Science and Technology (SS1320ES) and the University of Oxford (# 568-21) approved the study. Both face-to-face and virtual interviews were conducted privately after participants provided written consent; participants were compensated with 14 USD. Funding for this work was provided by the Wellcome Trust [215091/Z/18/Z].

## RESULTS

Forty-three participants, comprising 23 general medical personnel [i.e., nurses and doctors (MP)], nine consultant physicians and surveillance officers from regional referral hospitals (CP), five MoH case management pillar members (CM), five members of MoH top management (TM) and one WHO country office official, participated in the interviews (see Supplementary Files).

### Themes in the Interviews

Using thematic network analysis, we identified 58 thematic codes, of which 26 were subordinate to others (Level 2) (Supplemental files). Visual exploration (using ForceAtlas 2) was used to analyse the interactions between the 58 codes [8,37–41]. Figure 1 shows the relationships of codes to each other, visualising the average number of shared references with other codes (size of nodes), how often particular codes are discussed alongside each other in one reference (distance between codes and thickness of links), the total number of references shared between all codes (centrality to the network) and which codes exhibit closer relationships with each other compared to the rest of the codes (modularity). Modularity calculation identified five thematic topic clusters in the interviews: CMG Content & Updates (light green, 26.32%), Pandemic Preparedness and Response (violet, 26.32%), CMG Dissemination and Training (blue, 22.81%), Resourcing, Utilisation & Healthcare disparities (orange, 17.54%) and lastly, Patient Outcomes (dark green, 7.02%).

**Figure 1:**
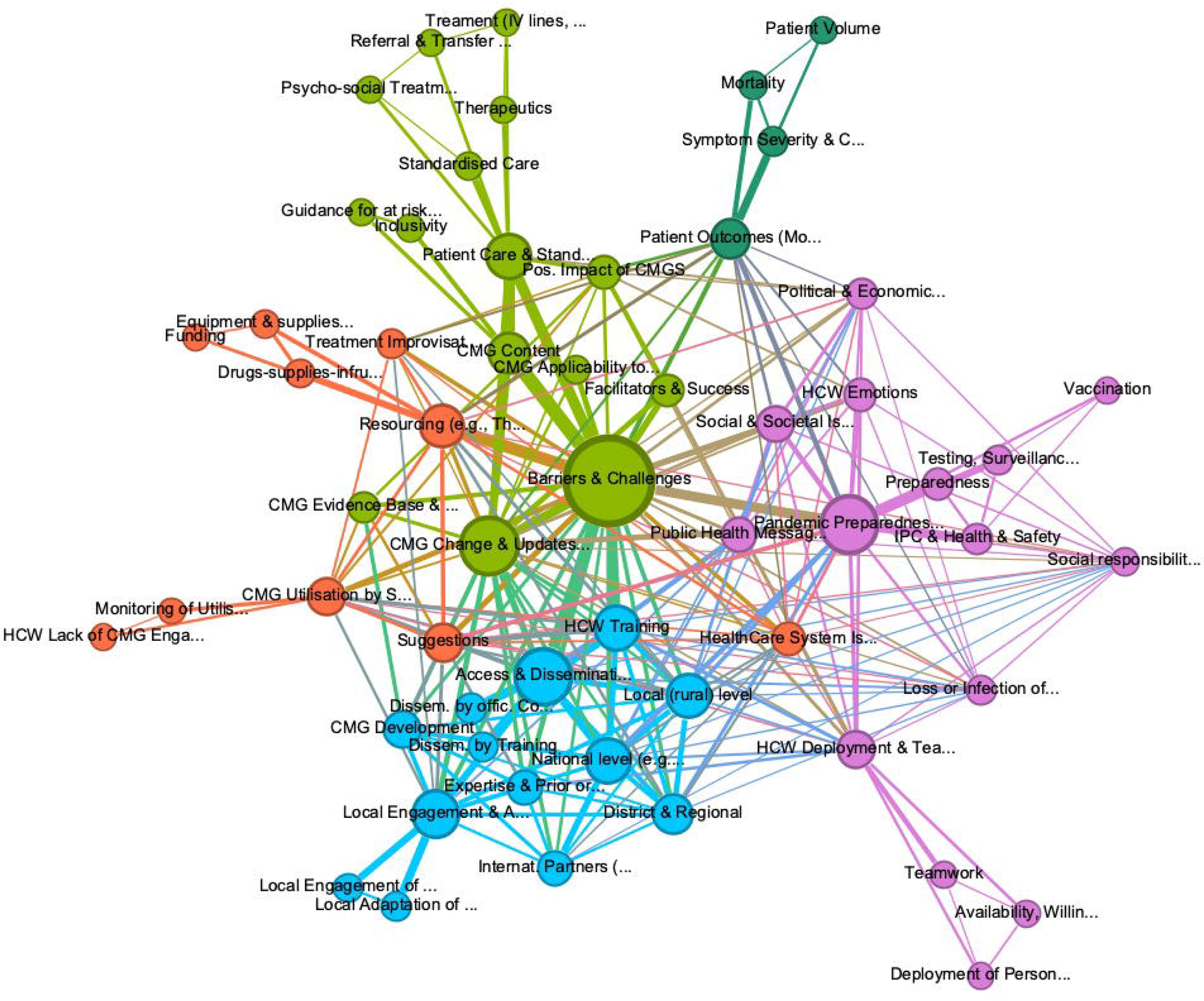
Graph of connections between all thematic codes, showing 58 codes, with 5 community clusters: CMG Content & Updates (light green), Pandemic Preparedness and Response (violet), CMG Dissemination and Training (blue), Resourcing, Utilisation & Healthcare Disparities (orange), Patient Outcomes (dark green). Lift >1.2, Graph Density: 0.154, Average Weighted Degree: 273.54, and modularity: 0.333.

The following sections will examine these five thematic topics in turn to identify the most frequently discussed barriers for the implementation of CMGs in Uganda and identify key suggestions by participants to overcome these.

#### CMG Content & Updating

A range of barriers related to CMG content and updates were discussed (e.g., standardization of patient care, guideline availability and updating, inclusivity of guidelines and difficulties adapting guidelines to local settings). The importance of CMGs for standardisation was frequently highlighted to reduce negative patient outcomes, increase shared understanding, reduce HCP anxiety about infectious disease work and equalise “*varied levels of expertise [and] not all units [being] equally equipped” (CM6).* However, CMG implementation appeared undermined by the applicability of guidelines and the attempt to standardise care across Uganda. Many participants therefore shared a sense that while guidelines developed for high income countries were difficult to convert to resource-constrained healthcare facilities, Ugandan CMGs were developed with little input of local stakeholders able to adapt guidelines to available local resources, e.g., “*if we are to have guidelines which can be useful, then we should tailor them to the local context of what is happening in Uganda*” (MP4) and “*our local teams [need to be] involved because they are the ones who are managing these outbreaks*” (CM3). Reluctance to use CMGs if they are not applicable to their setting was repeatedly described as reason for improvising patient care in accordance with locally available resources.

Similarly, CMGs were not equally inclusive of patient groups, with COVID-19 guidelines – due to the large amount of evidence-based guidance from international bodies – perceived as both inclusive of different at-risk patient groups (e.g., HIV/immunocompromised, pregnant women, children, elderly) and frequently updated to incorporate advances in changing evidence on pathogens and therapeutics. In contrast, CMGs for FVDs were reported as requiring more inclusivity:

> *“In West Africa [outbreak], we tried to develop guidelines for children. […] but the guidelines we were using were not sufficient to cover vulnerable groups especially children, pregnant women, elderly and people with disabilities. So, we were just cramping everyone together” (MP9).*

Many participants felt therefore that too much emphasis was put on international research in CMG development and not enough inclusion of local evidence, despite local personnel often encountering a changing case presentation first.

> *“They used to call it the Viral Haemorrhagic Fever, because there was a lot of people either bleeding, or passing out blood in stool, or they are vomiting blood but when it came to the West African Ebola, we could not see much, and […so] we [now] call it Ebola Virus Disease (EVD)” (TM1)*.

Participants routinely emphasised the importance of CMGs being guided by existing evidence and research collated in Uganda, which would not only enable rapid integration of new knowledge regarding symptomatology or treatments into existing guidance, but also lead to increased utilisation of CMGs by staff (see Figure 1).

#### Pandemic Preparedness & Response

Participants frequently discussed overall pandemic preparedness and response, highlighting the importance of collaboration across pillars and the inclusion of the community to increase preparedness, e.g., *“When there is an outbreak, all pillars work together to respond; there is a lot of coordination and working together as a team”* (TM3). Nevertheless, logistical issues around surveillance and testing were discussed. For example, while surveillance infrastructure was better for COVID-19, lack of laboratories able to process FVD samples located at national level reportedly resulted in frequent diagnostic delays, e.g., *“for Ebola we could not even recognize until someone takes another few days to take the samples to Kampala, […] we must have a system of taking samples from one point to another” (MP17)*.

Many identified the lack of adherence to guidelines in the population, which hinders the effective response to outbreaks. For example, “*[Because of] religion and cultural beliefs, you find the community refusing that there is Ebola, they refuse to come to hospital, they report late [and] health workers also believe that their religion doesn’t allow them to perform some rituals*” (TM3). Similarly, participants routinely described that lack of pandemic preparedness, reluctance to engage with scientific information, and low medical literacy in the population alongside widespread misperceptions, myths, conspiracy theories and stigma (e.g., ‘witchcraft’, ‘government harvesting organ’ or ‘killing people to access natural resources’) impeded their ability to implement guidelines and carry out their job. For example, “*we had to withdraw because […] they want to beat you [when] they see you taking the dead body, [saying] ‘you have killed our relative because you want money’* (MP3). Considering these issues, the lack of templates for public health messaging during outbreaks alongside CMGs, which enable HCP to integrate community leaders, patients’ relatives and the population were discussed as problematic.

> *“Updates would be for everyone, the case management team, the health workers who are not part of the case management, the village health teams, even the community to create awareness. If everybody is aware and such an outbreak comes again, everybody would be on standby and it wouldn’t take us or kill us the way it did previously” (MP5)*.

The overlap between this thematic topic is visualised in Figure 1, showing the connection of ‘Social & Societal factors’, ‘healthcare system’, ‘patient outcomes’, ‘public health messaging’, and ‘pandemic preparedness’.

#### CMG Dissemination and Training

Availability of CMGs was reportedly patchy, with more frequently occurring, or internationally prioritised, HCIDs (e.g., Ebola disease and COVID-19) being prioritised over less frequent HCIDs, e.g., *“For Marburg there was little apart from checking the internet, so […] when we were confronted […] I based [treatment] on the old knowledge [from] seven years earlier” (MP19).* Logistical factors (e.g., scarcity of print copies, limited internet access and limited training for HCP) were emphasised as limiting CMG implementation. Many described that HCP often were unaware of guidelines, describing challenges accessing guidelines, associated with slow and patchy dissemination and limited announcements, e.g., *“They have not yet shared them and all of a sudden you come and land on the book about the new guideline on the internet by chance”* (MP7). Rural healthcare facilities lacked printed copies and frequently could not access online sources, which reportedly curtailed effective communication across healthcare facilities, access to health records, dissemination of guidelines and attendance of virtual seminars.

> *“[In] lower-level facilities […] you rarely get the Filovirus guidelines […]. Dissemination is really a big problem, [especially as] the common venue where we have these outbreaks is more in the rural areas where the human being interfaces with the wild population and that’s where you’re going to get the lowest level of health centres. We tend to concentrate at the big facility level while this should be going down there, and from down there it goes upwards” (TM4)*.

Alongside dissemination problems and lack of internet access, lack of applied training in HCID care was discussed as a central issue, e.g., *“lack of knowledge [because] people are not fully trained and if trainings are not practical, they will never get skills” (MP21).* The lack of applied training on guidelines and dedicated platforms to access them were also attributed to colleagues seeking false information and consequently applying questionable treatments of COVID-19 with repurposed therapeutics (e.g., ivermectin; hydroxychloroquine) even after their use was warned against by the WHO, CDC and the Ugandan MoH guidelines.

> *“There should be practical sessions and continuous follow up of training outcomes […] if you just come for a week and train people, let’s say on how to manage EVD, tomorrow if someone is faced with a patient, they will fail to manage electrolyte balance. Not because they don’t know, but because they did not have a practical session, and no continuous mentorship” (MP4)*.

Lack of training was not only associated with adherence to guidelines and standards of care but also with heightened fear and anxiety, with many participants describing a reluctance to work on the frontline due to lack of training. Conversely, training was reported to increase confidence and self-efficacy, suggesting that this would lead to better implementation of CMGs, e.g., *“before training, I was worried; but […] the training really increases your level of confidence; it improves you and you perform well”* (MP22). Figure 1 and 2 illustrate that most participants discussed ‘Barriers’ alongside ‘HCP training’ and ‘Access & Dissemination’, and ‘Change and Update of CMGs’.

**Figure 2:**
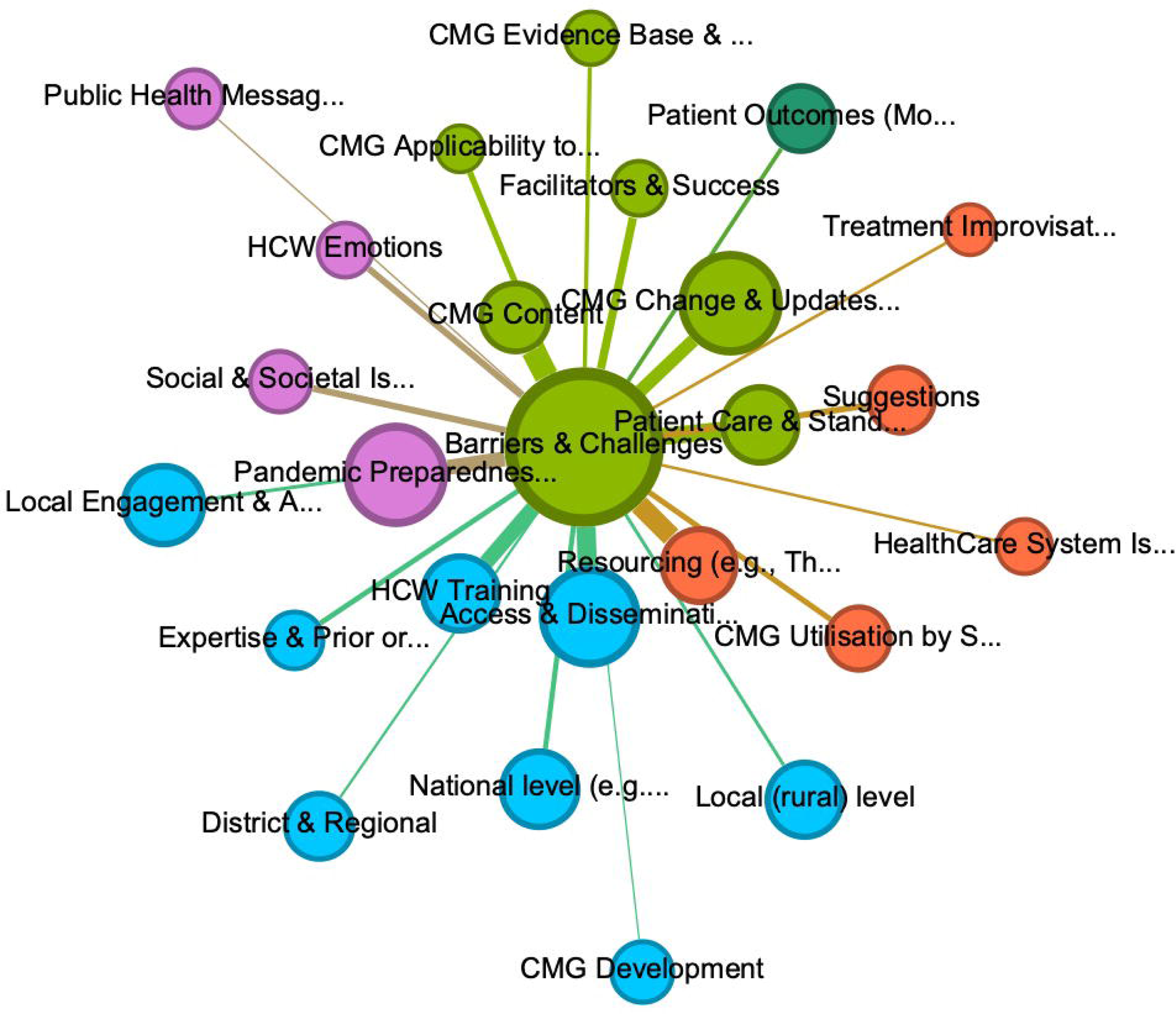
Overview of codes sharing most references with Barriers and challenges to the implementation of CMGs (adjacency and thickness of links). Lift <1.2 Edgeweight< 10; Fruchterman layout.

#### Healthcare Disparities, resource constraints, and limited utilisation

Problems with resourcing and utilisation were often discussed alongside issues involving the Ugandan healthcare system. Many highlighted the urban/ rural divide in the healthcare system, whereby severe cases are transferred to dedicated regional hospitals able to monitor oxygen, fluid and electrolyte balance, dialysis, etc., while lower-level rural facilities had none of these.

> *“When it comes to urban versus rural though, there are some things that apply in urban setting that may not apply in the rural setting, e.g., a patient who develops a complication while at home, can easily be evacuated to the facility in an urban setting, but in the rural setting, means of transport are so scarce that it may not work there” (MP2)*.

Participants acknowledged a discrepancy between nationally adopted guidelines and the regulatory and logistical realities on the ground, which limit universal CMG implementation, e.g., *“having guidelines [is one thing] but having the resources to buy what the guidelines recommend is another one”* (TM2). The universal implementation of CMGs reportedly is further inhibited by differences between private and public healthcare facilities, where private facilities may distribute therapeutics more quickly or instate different infection prevention and control (IPC) measures.

*In the private sector, there was a lot of resistance […] we discovered that there were some [expensive] drugs that had not been recommended to treat COVID 19, but they were using it and not adhering to our guidelines […]. We also had instances where clinics were reusing PPE, coveralls which were supposed to be single uses […they] sprayed it with chlorine on the outside and put it out to dry and then reused it.” (TM2)*.

Lack of resources was recurrently associated with increased infection risk and mortality, e.g., *“if the person doesn’t have masks, they won’t be safe, […if] they go and operate an Ebola case – like what happened in Mubende [in Nov 2022] – then they get infected” (TM3).* Similarly, lack of HCP posed a barrier for the implementation of CMGs, leading to issues in providing supportive care and upholding IPC protocols.

> *“One [issue] is human resources, if people are many, guidelines can be followed to the dot, but if people are few, certain things are forgotten, e.g., they say you’re not supposed to spend [more than] one hour in the PPE when you enter the ETU, but [if] you have 20 patients […] you end [up] extending [and] risking yourself (MP21)*.

For many, HCP shortage was most frequently attributed to lack of knowledge, experience, or preparedness, leading to unwillingness of HCP to work on the frontline during outbreaks, e.g., “*they fear to go and attack such a deadly disease without such knowledge*” (CP7). The intersection between access to guidelines, lack of resources and HCP’s ability to adhere to guidelines was most evocative in this reflection:

> *“Implementation is a different thing [as] the health workers knew what to do, [if] they didn’t do, it was because of lack of adequate infrastructure; particularly for critical care, [it was] an issue both in terms of human expertise and also infrastructure” (CM6)*.

Nevertheless, participants in more senior positions highlighted that existing guidelines and resources are often insufficient for improving CMG adherence, e.g., *“I […] found a nurse drawing blood with bare hands, no gloves [though] on the trolley were gloves, [so…] we have these guidelines but for some reasons they are not being followed [and] that’s why health workers in ETU and CTUs die”* (TM5). Several participants attributed this to a general *“poor reading culture – in Uganda we don’t read” (MP21)*, which may result in attempts to improvise care, e.g., “*if they don’t read on their own, they won’t know what to do, they will try to manage on their own and then get infected*” (TM2). However, others associated non-adherence with a lack of understanding of “*how serious the condition is […and] the availability of […] tools and other supplies, which also affects compliance” (CP3), and a general lack of monitoring, e.g., “there was no monitoring, there was no re-enforcement, so if Dr xx is using chloroquine as a treatment of COVID-19, what system will pick him or remand him?”* (TM5). Figure 1 visualises this intersection of resourcing, utilisation, monitoring and existing evidence base.

#### Patient Outcomes

CMGs were regarded as important to increase shared understanding and standardisation of care, and to reduce negative patient outcomes, as well as risk and anxiety about working with highly contagious patients. Nevertheless, some described that due to existing healthcare disparities and shortage of staff trained in HCID care, prioritizing one disease inevitably meant delivery of other essential services were impacted, e.g., *“HIV intensive care […], malaria, everything was affected because of one disease, so it’s more than important to prioritize these diseases […] and make sure that the health system is resilient enough to continue to support the other essential health care services”* (CM3). Some participants also highlighted indirect positive effects of adherence to CMGs, including reducing stigma and mistrust and increasing the populations adherence to guidelines:

> *“Patients that are managed according to protocols and guidelines have a better chance of survival and are being discharged to go home, [and] the more people that go back home, the more the community trust[s] us and the earlier the patients – if they […] get VHF – will come to treatment centres for testing and further management.” (MP20)*

The link of these positive outcomes of CMGs to *‘patient outcomes’* is visible in Figure 1.

Overall, Figure 2 (using a Fruchterman-Reingold layout algorithm [37–40]) highlights the codes most frequently discussed alongside barriers and challenges for the implementation of CMGs. As participants most frequently discussed CMG implementation to be hindered by ‘*Content of CMGs’* (e.g., lack of inclusion of at-risk patient groups and adaptation to local settings)*, ‘Patient Care & Standardisation’* (e.g., treatment, therapeutics, patient triage), *‘Resourcing’* (e.g., availability of therapeutics, equipment and personnel), *‘HCW Training’* (e.g., lack of training in CMGs*), ‘Access & Dissemination’* (e.g., lack of internet or print copies in rural areas), *‘CMG Change & Updates’ (e.g., slow updating for HCIDs) and ‘Pandemic Preparedness’ (in particular lack thereof).* Other codes (e.g., ‘*applicability to settings’, ‘healthcare system issues’*, *‘patient outcomes’,* and ‘*improvisation of treatment’* or ‘*utilisation of CMGs by staff’*) were also discussed but, as indicated through their distance, less frequently.

### Suggestions for the implementation of CMGs for HCIDs

Considering these issues, Figure 3 illustrates the participants’ suggestions to facilitate better implementation of CMGs, in particular training of healthcare staff alongside regular updating and dissemination, improving scope and inclusivity and engaging local stakeholders in the development process. The range of suggestions to increase CMGs in Uganda were best summarised by one of the interviewed doctors who had been working in a remote district during COVID-19:

**Figure 3:**
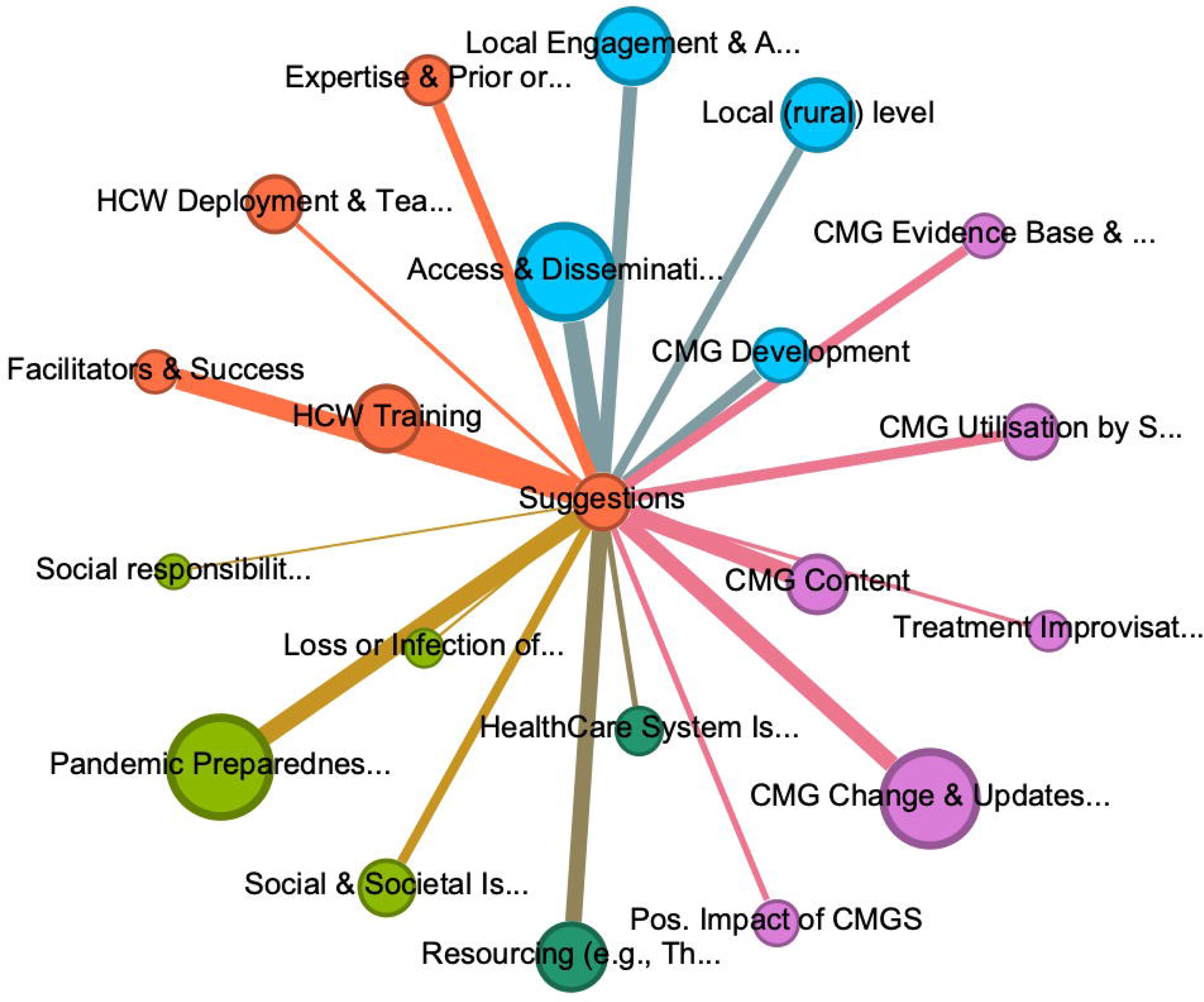
Thematic codes most frequently associated with suggestions for the implementation of CMGs. Lift <1, Edgeweight< 3, Fruchterman Layout

> *“First, all the stakeholders who develop guidelines should resist from copy and paste […], so, we read the guidelines [and] tailor them to what works for us. Second, [involve] the people who are managing patients on the ground, […] no matter the qualification. Even if someone is an enrolled nurse involved in the day-to-day management of patients, they practically know more than the professor who will pass there once in two months. Third, when these guidelines are passed on, it should be for practical purposes not for accountability purposes”. (MP13)*.

Cognisant of the difficulties of providing adequate healthcare resources in LMICs, participants recommended guidelines to be adapted to reflect local resource situations, while aligning procurement of therapeutics and equipment as well as surveillance with the suggested guidelines. Involvement of local stakeholders in CMG development was routinely debated to increase applicability to the local context, ensure quick reaction to new outbreaks and include at-risk populations (e.g., children, elderly, immunocompromised, pregnant women). While participants highlighted the necessary collaborations between international partners, medical-regulatory bodies, MoH and local stakeholders to ensure quick revisions with up-to-date evidence base for pathogens and therapeutics, they also emphasised the need for more local research efforts into HCIDs, which would increase HCP buy-in and ensure the applicability to the LMIC context.

Almost all participants described suggestions for better dissemination of CMGs to support the uptake and implementation of guidelines by HCP. These suggestions ranged from (a) improved internet access alongside living guidelines [16] to allow virtual, real-time access to guidelines, integrated patient health records, and communication; (b) dissemination of print copies, pamphlets, posters, and training materials into the facilities and (c) hands-on practical training of HCPs on guidelines. As such, participants acknowledged that internet access by itself would not increase implementation of CMGs but would require designated portals, email communication and social media campaigns to ensure rapid pervasive information dissemination of new updates. To ensure harmonization of training and standardisation of care across the healthcare system and increased educational opportunities for staff in rural healthcare facilities with less resources, most participants highlighted the benefits of cascading hands-on training with on-site local personnel in charge of training and championing the CMG. Using rehearsals and simulations, provided by experts experienced in managing HCIDs was linked to implementation of and compliance to CMGs.

Considering limited pandemic preparedness, the requirement for early capacity building and continued clinical education in infectious diseases (ID) skills to adequately prepare for an outbreak and counteract skill loss and staff outflow was highlighted, e.g., *“capacity building should be continuous for [IDs] because even those that had been trained retired [or] have gone to other sectors and others have forgotten” (CP4)*. As most outbreaks of FVDs were deemed to be predominantly rural, some participants suggested the utilisation of mobile lab equipment and personnel to facilitate faster testing and community surveillance and the deployment of a cadre of readily available response personnel to support and train local health facility staff.

Lastly, participants routinely discussed other aspects related to successful implementation of CMGs, which were not directly related to clinical management but rather to the inclusion of public health messaging and attempts to decrease myths and misperceptions, that may impact HCP willingness to work on HCID units and ensure compliance with guidelines during outbreaks. Table 2 provides an overview of concrete recommendations and suggestions from this research.

### Key Recommendations

**Table 2:** Key Recommendations for CMG Implementation in Uganda.

| Recommendations for CMG Updates, Dissemination, and Monitoring: |  |
| --- | --- |
|  | Include practitioners and local stakeholders in CMG development to ensure representation of views of personnel in rural and resource-constrained settings. |
|  | Align guidelines with locally available resources such as medications, equipment and supplies for better management outcomes. |
|  | Review and update FVD CMGs to include new evidence and management options available from clinical trials. |
|  | Increase access to guidelines through provision of living guidance, hard copies, posters, webinars and local facility CMG champions. |
|  | Integrate local research sites into HCID trials and incorporate local stakeholders to increase buy-in and adaptability to context. |
|  | Monitor CMG utilisation and evaluation of standards of care. |
| Recommendation for HCP: |  |
|  | Deliver practical training on HCIDs to HCP throughout their educational and professional development with periodic follow up to counteract reduced willingness of HCP to work in epidemic/pandemic response. |
|  | Utilize mentorships and cascading training (delivered by experienced personnel) to model clinical care, monitor compliance and increase self-efficacy and HCP preparedness. |
|  | Provide training on CMGs that is hands-on, practical, and skills-based, utilising rehearsals and simulations. |
| Resourcing of supportive care and research |  |
|  | Create readily deployable response teams to bolster and support local health facility staff and involve more physicians in outbreak response. |
|  | Resource lower-level facilities with mobile lab equipment and personnel to facilitate faster testing and community surveillance. |
|  | Align procurement of therapeutics and equipment and staff allocation with the guidelines by integrating them into the guideline development process. |
| Recommendations for Communities Outreach and Public Health Messaging: |  |
|  | Develop and invest in public health messaging alongside CMG development to address low levels of healthcare utilization, stigma and willingness of HCP to work. |
|  | Engage local communities to collaborate with HCP (e.g., village health teams) on case identification, psycho-social aspects of reuniting discharged patients and reducing stigma in the population. |
|  | Improve communication and coordination between different partners involved in outbreak response, including continuous updates of medical staff on new information and trends through virtual and non-virtual channels. |

## DISCUSSION

Previous research during the 2013-2016 EVD outbreak in West Africa has demonstrated the importance of optimised supportive care, whereby case fatality ratio among patients treated in settings providing supportive care with technologies for continuous physiological monitoring (oxygen, fluid and electrolyte balance, dialysis, monitoring, etc.) were lower when compared to patients treated in resource-constrained settings with limited access to technologies [42,43]. While many of our participants made similar claims, the presented findings provide evidence that CMGs for the provision of standardised care are often the first and only defence against an outbreak and illustrate some of the key conceptual, logistical, and practical barriers inhibiting the implementation of CMGs in LMIC contexts.

Concretely, the thematic analysis supplemented by social network analysis highlights that CMG implementation in Uganda is hindered by five concerns regarding: (1) deficient content and slow updates of CMGs (i.e., slow, exclusive updating process too reliant on international evidence and thus not applicable to local setting and limited inclusivity of at risk-populations); (2) pandemic preparedness and response (i.e., lack of surveillance and testing infrastructure, and stigma and misperceptions in the population impacting the implementation of CMGs by HCP); (3) slow dissemination and lack of training (i.e., lack of print copies, internet access and dedicated hands-on training on standards of care); (4) scarce resources and healthcare disparities which impact the utilisation of CMGs (i.e., rural/ urban differences in healthcare sector, alongside incomplete monitoring hinders the implementation of CMGs); (5) patient outcomes (i.e., prioritisation of HCID leads to restricted delivery of services).

Many of our results were reminiscent of previous findings, highlighting the limited awareness of existing guidelines by HCP [26], limited access to treatments and equipment [1], as well as limitations of availability, inclusivity, and applicability of HCID CMGs across global LMIC contexts [1–6]. Nevertheless, besides providing key recommendations for the improvement of CMG implementation from HCP involved in the most recent HCID outbreaks (e.g., COVID-19 and SVD in 2022), our results go beyond previously reported findings, by emphasising the complex relationship between many of the conceptual, logistical, and practical factors which hinder or facilitate CMG development and implementation. This study was able to display how insufficient training, patchy dissemination and slow updating exacerbate many of the underlying difficulties in LMIC contexts, by illustrating how these issues are related to resource constraints, healthcare disparities and limited surveillance and referral infrastructure. While CMGs can inform efforts to improve equity in access to best available evidence-based care [14,15], without addressing some of the presented underlying causes of limitations to availability, accessibility and applicability, HCP will not be able to implement the standards of care necessary to quell HCID outbreaks. For example, investment in internet access at local hospitals, developing living guidance and integrating local stakeholders in the CMG development process can help to address local exigencies, resource constraints and misperceptions that limit the successful implementation and utilisation of CMGs by HCP.

While this case study of Uganda is not generalizable to a global context, the findings nevertheless hold value for other LMIC contexts. For one, the serendipitous inclusion of participants deployed during the 2022 SVD outbreak in Uganda provides evidence from the most recent HCID outbreak. Through the focus on HCP with considerable working experience on COVID-19 or FVD Treatment Units in Uganda, our study was able to collate a range of concrete and actionable recommendations from the interviews, which may be illustrative for those seeking to develop practical CMGs for supportive care in HCID outbreaks. By focusing greater attention on the realities of guideline adoption and implementation, insights from this work can not only inform other countries’ responses to HCID epidemics but also provide recommendations for supranational, governmental and non-governmental organisations advising and funding CMG development. Moreover, this work will inform implementation of supportive care guidelines in future HCID outbreak scenarios where evidence for clinical management may be initially limited.

## CONCLUSION

This in-depth case study of Uganda aimed to explore contextual and supplementary factors which hinder or facilitate CMG development and implementation in Uganda and underlines the significant obstacles to implementing CMGs in LMIC contexts, whereby outdated guidelines, poor dissemination and training deficits exacerbate prior healthcare disparities and resource constraints. For example, otherwise small issues (e.g., lack of internet access, vehicles, misinformation) can compound to have wider consequences for the implementation of CMGs by limiting access to training, communication, diagnostics and impact the populations behaviour and HCP willingness to work on HCID units. This study emphasised that CMG development in LMICs needs to extend beyond adaptation of international clinical evidence for HCID treatment but requires adjusting for the available local epidemic and pandemic preparedness and response capabilities and demands. Besides highlighting barriers to CMG implementation, the research also offers actionable recommendations to enhance CMG implementation, which underscore the necessity of integrating local stakeholders to ensure guidelines are reflective of the reality of the local health system, applicable and inclusive of resource-constrained settings, disseminated widely and in a living manner and complemented by cascading hands-on training. While these findings from Uganda are not universally applicable, they offer valuable insights for LMICs to improve HCID outbreak responses and for organizations involved in guideline development and funding.

## Supporting information

Interview Guide Health care workers

Interview Guide Policy Makers

COREQ Checklist

Participant Information Table

## SUPPLEMENTARY DATA

### Data availability statement

The datasets generated and analysed for this study can be found at https://osf.io/djv52/?view_only=e8139fa28c404285829b56c638bf7f47

### Patient consent for publication

No patients were involved in this study; however, preliminary results informed subsequent research by the ESHCID team focusing on patient experiences during HCID outbreaks in Ebola, Uganda and Sierra Leone.

---

Table 3: Overview of HCID outbreaks in Uganda with data from [27–30]

Table 4: Key Recommendations for CMG Implementation in Uganda

