## Supplementary material for "‘*Involve those, who are managing these outbreaks*’– Identifying barriers and facilitators to the implementation of clinical management guidelines for High-Consequence Infectious Diseases in Uganda": Interview Guide Health care workers

walimu

Version 2.0 17 October, 2022

**APPENDIX 6: SEMI-STRUCTURED INTERVIEW-LOCAL HEALTH STAFF**

**Study Title: Evaluation of Supportive Care Guidelines for Epidemics: Implementation Case Study—Filovirus Diseases (FVDs) and Covid-19 Guidelines in Uganda.**

Time required: 30 minutes – 1 hour

**Nurses, Doctors, physicians and surveillance officers involved in responding to Ebola outbreak.**

**EEXPERIENCE IN EBOLA RESPONSE**

- Tell me about your role in the current Ebola (Sudan virus disease SUDV) outbreak
- Could you please tell me about the current Ebola SUDV outbreak? How have you previously participated in responding to similar outbreaks? If yes when and how? Personal experiences Narrative stories
- Did you or your colleagues feel prepared at the beginning of the epidemic or when called upon to respond to Ebola SUD? Why or why not?

**Personal Factors (Factors Related to Respondent's Knowledge and Attitude to Management of Emergency Transmission Diseases In their Community)**

- What do you think is the severity of **Ebola SUDV** infection and what is the situation of in community?
- how did you get information about this current outbreak? (from colleagues, hospitals, internet, etc.) Do you think there is a need for an update on Ebola management for medical staff?
- Do you feel confident in treating patients with Ebola SUDV? Why or why not? What could help you feel more confident in how to treat patients with Ebola SUDV in the current outbreak?

**A. APPROPRIATE AND UPDATED CLINICAL MANAGEMENT GUIDELINES, SUPPORTIVE TREATMENT AND TREATMENT RECOMMENDATIONS**

- Are there currently available clinical management guidelines for the diagnosis and treatment of Ebola SUV? (international/national/regional/hospital/collegiate guidelines etc.) if yes, which ones?
- Are you aware of that the WHO newly released EVD therapeutic guideline for Zaire ebolavirus in Aug. 2022, which is designed as an "add-on" to their 2019 EVD guideline on supportive care?
- Were you familiar with these guidelines before the outbreak? Are these guidelines applicable for all Ebola viruses (e.g. Zaire virus and SUDV)? How are the current guidelines working compared to how they used them before?

**B. IF AVAILABLE EBOLA CLINICAL MANAGEMENT GUIDELINES:**

***Knowledge of Ebola clinical management guideline,***

- In your understanding, who was involved in its preparation?
- How are the guidelines communicated to clinicians? (e.g. through training/education sessions/finding the right moment/requires personal effort/not communicated) via other staff
- Do the guidelines which you know cover the elderly population, children, pregnant women, those living with HIV? Have the guidelines been developed based on evidence?
- Do you feel these guidelines are up to date? If yes or no, why? If yes, how were these guidelines updated?



MAKERERE UNIVERSITY
SCHOOL OF PUBLIC HEALTH
APPROVED
VALID UNTIL
20 JUL 2023
RESEARCH & ETHICS COMMITTEE
P O. BOX 7072, KAMPALA

- Are the guidelines applicable to or could these guidelines be adapted for different contexts? (e.g. different institutions, geographic areas, urban vs. rural, different resources and/or technology, humanitarian crises)
- So far, have there been any training on the use of these guidelines. If yes, Which one in particular? Was the training effective? What still needs to be added to the training? Based on the training, do you feel confident and well prepared to respond?
- Do you have access to the supportive care (e.g. fluids, monitoring of electrolytes, glucose etc) recommended for patients in the guidelines to enable implementing the recommendations/treat your patients with these? Yes, No If no, please explain what is needed.

**Experience with Recommended Diagnostic and Treatment Guidelines and Factors Which Support or Hinder Their Use**

- Do you feel confident about the correctness of the contents in available guidelines? Do you believe in the method which suggested? Do you have any concerns when using the guidelines?
- Are there any issues that have not been covered in that guidelines? If so, what things are missing and need to be added to the guidelines? What do you do when you find a case?
- Do you feel well protected and safe to be able to protect others? If yes or no, what makes you feel so?
- So far, what has been the final condition of patients (outcome) when treated using the guidelines? What do you think went wrong or needs to be included to improve patient outcomes?
- What are the factors might currently support the use of /implementation of the treatment recommendations in these guidelines by health workers? What might hinder their use?

Knowledge and skills (are there continuing education/professional development available or not?)

- Hospital resources (including e.g. staff, oxygen, fluids, symptomatic treatments, electrolyte monitoring equipment, isolation rooms/beds)
- Support for you (staff/educational/clinical/resources/management)
- Contextual factors (culture/community/resources/internal, institutional, state, local politics/medical education)

- Are you involved in the development and implementation of guidelines for the diagnosis and treatment of other infectious diseases, such as **Ebola,** SUDV and Marburg and COVID-19? If so, how does that experience compare to your experience of treating patients with **these diseases?**

**Perceived benefits of using Ebola clinical management guidelines:**

- What do you think are the benefits of using these guidelines for patients and their impact on public health? How do you know the patient outcome was as a result of using the guidelines?
- Development of clinical guidelines for research: Have you ever conducted research using these guidelines? So far, how have these guidelines been used in a study?)
  - Research implementation
  - Impact on final condition of patient (outcome) - (Was there a difference when using or not using the guidelines in the case Ebola?) What are some examples you know of?
- If no, do you think clinical management guidelines are also needed for research and development efforts related to FVDs and/Covid-19?

**C. IF FVDs AND/COVID-19 CLINICAL MANAGEMENT GUIDELINES ARE NOT AVAILABLE:**

- what do you base your treatment decisions on? (previous knowledge, text books, colleague's /superior's advice, senior management advice, other – please describe)
- Who do you think should be involved in the process of decision making, development and implementation of the **FVDs** and/Covid-19 clinical management guidelines?

**D. Lessons learnt and Recommendations**

- In your opinion, what are the key lessons learnt from in using the guidelines in this particular outbreak?
- What are your top recommendations for the development of Ebola guidelines and improving the overall response of EVD now and in the future?
  - E.g how to improve coordination
  - Resources

MAKERERE UNIVERSITY
SCHOOL OF PUBLIC HEALTH
APPROVED
VALID UNTIL
★ 20 JUL 2023 ★
RESEARCH & ETHICS COMMITTEE
P O. BOX 7072, KAMPALA
