## Supplementary material for "‘*Involve those, who are managing these outbreaks*’– Identifying barriers and facilitators to the implementation of clinical management guidelines for High-Consequence Infectious Diseases in Uganda": Interview Guide Policy Makers

### APPENDIX 5: SEMI-STRUCTURED INTERVIEWS –POLICY MAKERS AND GUIDELINE DEVELOPERS

**Study Title: Evaluation of Supportive Care Guidelines for Epidemics: Implementation Case Study— Filovirus Diseases (FVHDs) and COVID-19 Guidelines in Uganda.**
**Interview guideline for Ministry of Health/WHO/CDC,**

Are there currently available clinical management guidelines for the diagnosis and treatment of FVDs (Ebola and Marburg) and/Covid-19 in Uganda If yes to question 1, ask questions on section A and D. If no, skip to B and D.

**A. If yes,** which ones?

1. Are you aware of that the WHO newly released EVD therapeutic guideline for Zaire ebolavirus in Aug. 2022, which is designed as an "add-on" to their 2019 EVD guideline on supportive care?

2. In the current context of the (Ebola) Sudan virus disease outbreak (SUDV), do you have access to guidelines to guide clinical management for patients presenting with SUDV? If yes, which guidelines would you use?

3. In your understanding, who was involved in preparation of clinical guidelines for?

4. In your opinion, how is the severity of FVDs and/COVID-19 infection? and what is the situation of FVDs and/COVID-19 in the community? How important is the FVDs and/COVID-19 issue compared to other infectious disease issues, for example: dengue fever, HIV, etc.?

5. How are the available clinical management guidelines communicated to clinicians? (e.g. through training/education sessions etc.)

6. Is the treatment recommendations in these guidelines sufficient? Do they cover for the whole population? cover the elderly population, children, pregnant women, and those living with HIV? Have the guidelines been developed based on evidence? Are they of good quality? Is there any other information you wish the guidelines included?

7. Could the guidelines be used in or adapted to be used in different contexts? (e.g. different institutions (community care, hospitals), geographic areas, urban vs. rural, different resources and/or technology, humanitarian crises)

8. In your understanding, what influenced the adoption and the use of these clinical guidelines?

9. Is implementation of SUDV clinical management guideline recommendations in healthcare centres supported by a review of need for:
   - Healthcare worker training need
   - Resources (e.g. fluids, monitoring of electrolytes, glucose etc)
   - others: please explain
10. In your understanding, what are the factors that influence the decision in the preparation and development of the FVDs and/COVID-19 clinical management guidelines? What is the decision-making process for approving the guidelines? Collegial or just one-sided?
11. Have the guidelines been updated continuously? In your understanding, how was the guideline updated and what were the factors that influenced the change?

MAKERERE UNIVERSITY
SCHOOL OF PUBLIC HEALTH
APPROVED
VALID UNTIL
★ 20 JUL 2023 ★
RESEARCH & ETHICS COMMITTEE
P. O. BOX 7072, KAMPALA

12. How do you think the availability and quality of clinical guidelines impact the outcomes of patient clinical management, and final patient outcomes? How to monitor and evaluate the use of this guide with treatment outcomes/outcomes?
13. Do you perceive that having access to standardised evidence-based clinical management guidelines have an impact on research and development efforts related to Ebola, SUDV and/COVID-19? (Pompt: e.g by standardising care to allow comparison with new interventions/treatments)
14. How have the CMGs impacted on how the community responds to patients after discharge?
15. What are the factors that you think might currently support the use of /implementation of the treatment recommendations in these guidelines by health workers? What might hinder their use? Are they aware that training is needed? And what resources might be needed?
16. Knowledge and skills (are there continuing education/professional development available or not?)
    - Hospital resources (including e.g. staff, oxygen, fluids, symptomatic treatments, electrolyte monitoring equipment, isolation rooms/beds)
    - Support for you (staff/educational/clinical/resources/management)
    - Contextual factors (culture/community/resources/internal, institutional, state, local politics/medical education)

**B. If there is no clinical management guideline for** FVDs **and/COVID-19 in Uganda:**

1. If no, what do you base your treatment decisions of patients with SUDV on?
2. In your opinion, how is the severity of FVDs and/COVID-19 infection? and what is the situation of FVDs and/COVID-19 in the community? How important is the Ebola, SUDV and/COVID-19 issue compared to other infectious disease issues, for example: dengue fever, HIV, etc.?
3. In your opinion, how important is the development of clinical management guidelines regarding FVDs including Sudan virus and/COVID-19 for medical staff? What are the benefits? Is there a demand or need for Ebola (SUDV) and/COVID-19 clinical management guidelines from medical staff?
4. In your understanding, what are the factors that hinder the development of FVDs and/COVID-19 clinical management guidelines to date? (politics, priorities, socio-cultural, funding, etc.) In your opinion, what are the ways to overcome these obstacles?
5. Who do you think should be involved in the decision making, development and implementation of FVDs and/COVID-19 clinical management guidelines?
6. In your opinion, what information should be included in the FVDs and/COVID-19 clinical management guidelines? (E.g. reflecting on the current Sudan virus outbreak)
7. If the guidelines have been developed, how should they be communicated to medical staff?
8. In your opinion, what are the FVDs and/COVID-19/s of the unavailability of clinical management guidelines on medical staff performance and patient outcomes?
9. Do you think clinical management guidelines are also needed for research and development efforts related to Ebola and/COVID-19?

**Lessons learnt and recommendations**

1. How do you feel Uganda, overall is responding to this outbreak using the available guidelines?
    - Preparedness at the beginning of the outbreak?
    - Strengths and weaknesses of the response
    - Clinical management of patients
    - Protection of healthcare workers

MAKERERE UNIVERSITY
SCHOOL OF PUBLIC HEALTH
APPROVED
VALID UNTIL
★ 20 JUL 2023 ★
RESEARCH & ETHICS COMMITTEE
P O. BOX 7072, KAMPALA

2. What are the key lessons learnt from this specific outbreak in relation to the clinical management guidelines? (availability, scope , supportive care and treatment recommendations, evidence base, quality)
3. Considering anything, what are your top recommendations for improving the overall response of EVD now and in the future?
   - E.g how to improve coordination
   - Resources
   -

MAKERERE UNIVERSITY
SCHOOL OF PUBLIC HEALTH
APPROVED
VALID UNTIL
20 JUL 2023
RESEARCH & ETHICS COMMITTEE
P. O. BOX 7072, KAMPALA
